# Automated speech analysis for risk detection of depression, anxiety, insomnia, and fatigue: Algorithm Development and Validation Study

**DOI:** 10.1101/2024.03.20.24304577

**Authors:** Rachid Riad, Martin Denais, Marc de Gennes, Adrien Lesage, Vincent Oustric, Xuan-Nga Cao, Stéphane Mouchabac, Alexis Bourla

## Abstract

**Background:** While speech analysis holds promise for mental health assessment, research often focuses on single symptoms, despite symptom co-occurrences and interactions. In addition, predictive models in mental health do not properly assess speech-based systems’ limitations, such as uncertainty, or fairness for a safe clinical deployment.

**Objective:** We investigated the predictive potential of mobile-collected speech data for detecting and estimating depression, anxiety, fatigue, and insomnia, focusing on other factors than mere accuracy, in the general population.

**Methods:** We included n=865 healthy adults and recorded their answers regarding their perceived mental and sleep states. We asked how they felt and if they had slept well lately. Clinically validated questionnaires measuring depression, anxiety, insomnia, and fatigue severity were also used. We developed a novel speech and machine learning pipeline involving voice activity detection, feature extraction, and model training. We automatically analyzed participants’ speech with a fully ML automatic pipeline to capture speech variability. Then, we modelled speech with pretrained deep learning models that were pre-trained on a large open free database and we selected the best one on the validation set. Based on the best speech modelling approach, we evaluated clinical threshold detection, individual score prediction, model uncertainty estimation, and performance fairness across demographics (age, sex, education). We employed a train-validation-test split for all evaluations: to develop our models, select the best ones and assess the generalizability of held-out data.

**Results:** The best model was WhisperM with a max pooling, and oversampling method. Our methods achieved good detection performance for all symptoms, depression (PHQ-9 AUC= 0.76F1=0.49, BDI AUC=0.78, F1=0,65), anxiety (GAD-7 F1=0.50, AUC=0.77) insomnia (AIS AUC=0.73, F1=0.62), and fatigue (MFI Total Score F1=0.88, AUC=0.68). These strengths were maintained for depression detection with BDI and Fatigue for abstention rates for uncertain cases (Risk-Coverage AUCs < 0.4). Individual symptom scores were predicted with good accuracy (Correlations were all significant, with Pearson strengths between 0.31 and 0.49). Fairness analysis revealed that models were consistent for sex (average Disparity Ratio (DR) = 0.86), to a lesser extent for education level (average Disparity Ratio (DR) = 0.47) and worse for age groups (average Disparity Ratio (DR) = 0.33).

**Conclusions:** This study demonstrates the potential of speech-based systems for multifaceted mental health assessment in the general population, not only for detecting clinical thresholds but also for estimating their severity. Addressing fairness and incorporating uncertainty estimation with selective classification are key contributions that can enhance the clinical utility and responsible implementation of such systems. This approach offers promise for more accurate and nuanced mental health assessments, benefiting both patients and clinicians.

## Introduction

Depression and anxiety disorders are recognised as the leading causes of disease burden [1], and their prevalences are high during the entire lifespan, across the sexes and all around the globe [2]. This burden was aggravated by the COVID-19 pandemic [3]. In these disorders, early identification and evaluation of severity of the symptoms are of prime importance since the incidence of suicide is associated with a diagnosis of depression more than 50% of the time [4]. Besides, measurement-based care, via the use of clinically valid scales, improves the follow-up and treatment of affected individuals with mental health disorders [5]. Timely interventions lead to better outcomes in mental health. This proactive approach can ensure early access to treatment and prevent significant complications. Yet, measuring mental health remains a challenge, since manifestations of depression and anxiety are heterogeneous [6], and co-occur with insomnia [7] and fatigue [8].

The exhaustive and objective assessment of these different mental health dimensions through validated assessment scales is long and fastidious for clinical staff, and particularly not adapted to primary care, which is at the forefront of handling mental health disorders[9]. The development of objective biomarkers, easy to collect without synchronization of clinicians and patients, has the potential to overcome these limitations. These quantifiable measures, encompassing biological, genetic, or behavioural assessments, could revolutionize early detection, enabling timely and targeted interventions that ultimately improve the patient’s outcomes and well-being. This is particularly significant for screening the general population across diverse mental health dimensions. Indeed, screening the first signs of mental health problems/symptoms could help to avoid escalation of symptoms, as different dimensions interact in time [7, 10].

The study of speech biomarkers in mental health holds great potential, offering a non-invasive and easily accessible avenue to capture significant motor, cognitive and behavioral changes due to mental health disorders such as depression [11–14]. Clinical evidence and research studies have increasingly linked specific automated extracted speech features, such as prosody, articulation, and fluency, with various mental health conditions, such as depression [11, 15], anxiety [16], suicide-risk assessment [17], fatigue [18, 19], or sleep deprivation [20]. The complexity of human speech extends beyond the intricate motor coordination involved. The speech production system within the brain relies on the synchronization of diverse cognitive, social, and motor processes [21, 22]. This intricate interplay involves hundreds of muscles across the respiratory, phonatory, and supralaryngeal systems, working in concert with critical cognitive skills like attention, memory, and planning. Additionally, social skills such as theory of mind and emotional processing play a vital role. Importantly, disruptions in any of the aforementionned motor, cognitive or social skills, as well as mental health states, can introduce perturbations in the resulting speech signal.

Besides, beyond research evidence, clinical practitioners also use voice (un)consciously when evaluating individuals, and these subjective evaluations could be complemented and refined with objective measures from automatic speech analysis.

Speech biomarkers emerge also as a promising avenue for mental health assessment due to their unique characteristics: they are non-invasive, cost-effective, and convenient tools. Recent hardware and software advancements have significantly simplified and reduced the cost of acquiring acoustic data, making it a more accessible option compared to traditional biological, imaging, or cognitive markers. In addition, speech data collection requires minimal effort from both patients and clinicians and can even be conducted remotely, further enhancing its feasibility in various settings.

However, despite its promises, the study of speech biomarkers remains largely fragmented, in laboratory settings, or not evaluated for deployment into clinical practice. This gap calls for more evidence to be integrated into clinical practice [23].

Research on speech in mental health in the general population often focuses on one isolated mental health dimension, even though there are proofs supporting the existence of networks of symptoms and syndromes in mental health that influence each other [24, 25]. Also, previous speech studies were limited to specific populations such as students [26] or the elderly[27]. However, machine learning models making predictions at the individual levels in medicine should be ‘fair’ and have the same quality of prediction no matter the demographics before they can be deployed in a clinical setting [28]. Finally, speech-based systems should not be tested only for the simple classification of binary labels (ex: depressed or not depressed), but rather for the estimation of the severity of symptoms [29] and their ability to refrain from giving an output when uncertainty is too high, therefore deferring decisions to the health staff in practice[30].

In this study, the main objective was to assess the predictive potential of speech models in detecting and estimating the severity of depression, anxiety, fatigue, and insomnia within the general population, using mobile-collected speech and mental health data. Besides, to prove that these models could be effectively implemented in diverse real-world settings, they are assessed for their fairness and uncertainty capabilities.

## Methods

### Participants

We recruited French healthy adult participants,, without any known severe psychiatric or neurological disorder (self-declaration), or speech pathologies such as stuttering or clutter.

All participants signed an informed consent form to participate in the study, in line with the Declaration of Helsinki, current Good Clinical Practice guidelines, and local laws and regulations. All procedures were approved by the French National Institutional Review Board, (identifier 23.00748.OOO2L7#I for the Committee for the Protection of Persons”).

### Study procedure

The participants completed the protocol on smartphones through the Callyope research mobile application in a home environment. The participants completed self-assessment scales for different mental health dimensions and recorded different speech tasks. In this work, we only focused on one spontaneous and semi-structured speech task, where participants had to answer: “Describe how you are feeling at the moment and how your nights’ sleep have been lately.” [31]. The participants were included by speech pathologist interns and recruited through social media platforms. Finally, we examined self-reported symptoms with clinically validated questionnaires (See Figure 1a.). Participants followed the instructions displayed on the Callyope application and their vocal answers were recorded with the smartphone’s microphone. The audio was sampled at 44.1 kHz with a 16-bit resolution. Each participant was asked to place his phone on a flat surface (for example a table) with the microphone pointing towards the speaker i.e. himself. The session should take place in a quiet environment, whenever possible.

**Figure 1.**
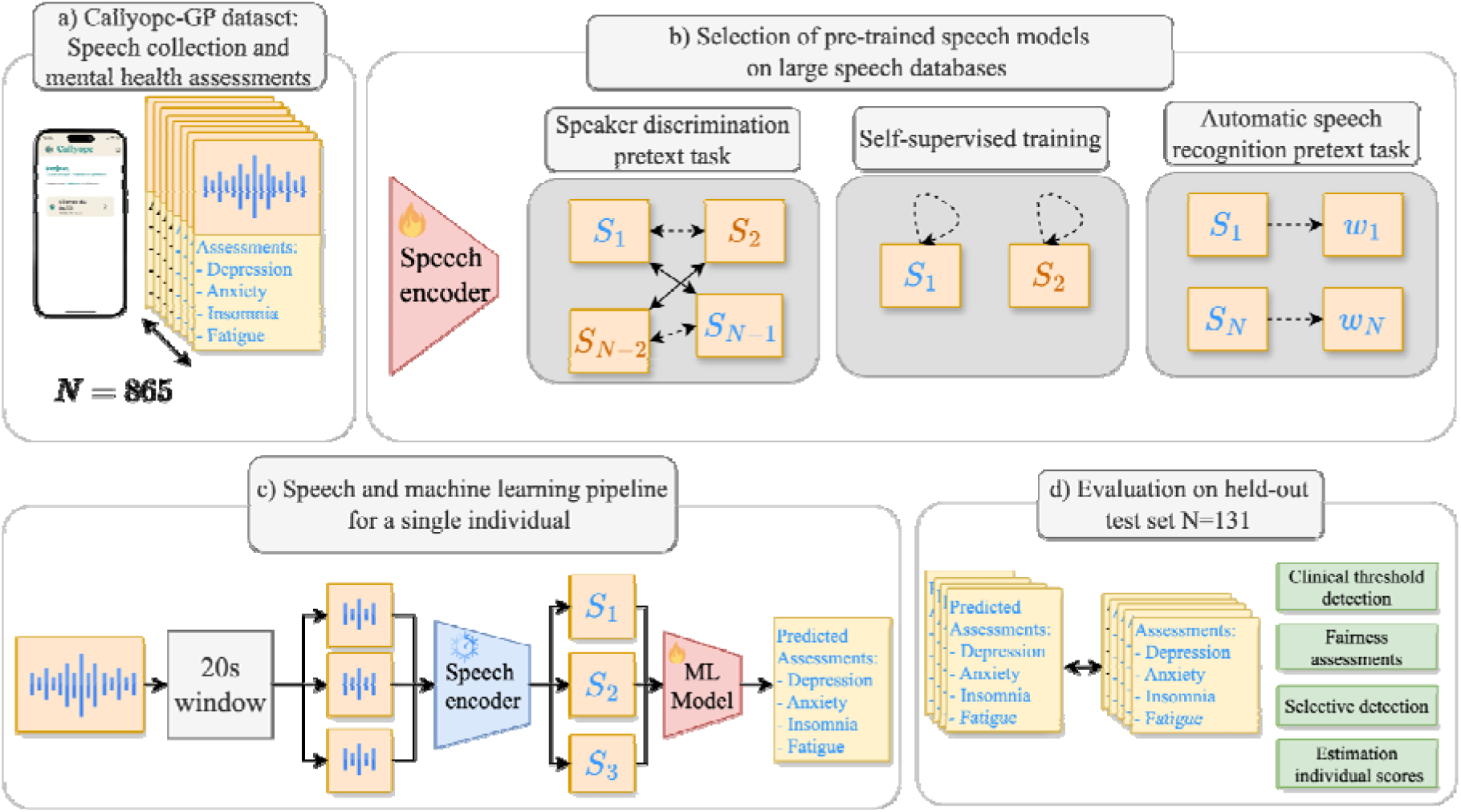
Overview of this study. (a) Overview of our Callyope-GP dataset, with 865 included participants (b) Flowchart of the pre-training phases of our speech encoders with an illustration of the pretraining speaker embedding process. (c) Graphical illustration of our speech and machine learning pipeline for a single individual. The pre-trained speaker embedding is frozen and only machine learning models on top are trained on training data. (d) Evaluation and comparison between true and predicted assessment on held-out participants (test set N=131). Symbols represent speech turn vector embeddings obtained from the speech encoder model. Colours represent speaker identities for each speech turn and embedding, represent the words spoken in the given audio.

We refer to the dataset collected in this study as the Callyope-GP dataset. We split randomly the Callyope-GP dataset into three sets: training, validation and testing. Demographic data, such as sex, age, and education level, were collected. We compared groups with adequate tests for their demographics and self-assessments to ensure that groups were consistent.

#### Measures of depressive symptoms

To allow broader use of our solutions, we assessed the severity of depression through the Beck Depression Inventory (BDI) [32] and Patient Health Questionnaire-9 (PHQ-9) [33] self-report questionnaires. In current clinical practices, it is common that different professionals interacting with a patient use different metrics to monitor depression. While depression assessment through these two measures exhibits a robust correlation at the group level [34], thus facilitating the development of an equational conversion for research uses, their limited efficacy at the individual level impedes their reliable conversions to predict individual depressive status [35].

The PHQ-9 is a short, self-administered questionnaire mainly used to screen and measure the severity of depression[33] and is sensitive to potential changes [34]. It includes the two cardinal signs of depression: anhedonia and depressed mood. We considered a risk of depression if the total score for the PHQ-9 was more than 10 (PHQ-9≥10).

The BDI is a self-administered questionnaire with 21 items, each centered around a core theme [32, 36]. Respondents are presented with statements for each item, and they are instructed to choose one statement, which is then associated with a score ranging from 0 to 3. The cumulative score for the scale can reach a maximum of 63 points. We considered the BDI threshold to be positive if the total score was more than 10 (BDI≥10), as it is above the normal range as defined by the authors of the BDI [36].

#### Measure of anxiety

The Generalized Anxiety Disorder questionnaire (GAD-7) is to measure or assess the severity of generalised anxiety disorder (GAD) [37].. This is a self-administered questionnaire that takes less than 5 minutes to complete, and it was especially developed to be deployed efficiently in primary care. The optimal cutoff for the GAD-7 was found to be a cut-off for the total score of GAD-7≥10 [37].

#### Measure of insomnia

The Athens Insomnia Scale (AIS) is a self-administered questionnaire to assess the patient’s sleep difficulties according to the ICD-10 criteria [38]. The AIS-8 comprises 8 items (5 minutes) and is a good tool for general sleep assessment and insomnia screening, and to measure the intensity of sleep-related problems, but also as a screening tool in reliably establishing the diagnosis of insomnia. The optimal cutoff for diagnosis to detect insomnia troubles, for the AIS scale, is 6 [39, 40].

#### Measures of fatigue

We used the Multidimensional Fatigue Inventory (MFI) to assess the different dimensions of fatigue [41–43]. It is a short self-report questionnaire (5-10 minutes) based on 20 questions to determine five dimensions of fatigue: general fatigue, physical fatigue, reduced motivation, reduced activity and mental fatigue. We also reported the total fatigue score as the sum of all sub-components.

We used the normative data from [42, 44] to choose thresholds for each sub-component. Individual sub-components of fatigue in the 75% quantile in the studied populations are all above 10. Therefore, we aimed to predict individuals’ scores which are above or equal to 10, for each dimension. As mentioned also in [42], the total score has clinical significance and validity, as it was observed to have the highest correlations with anxiety, depression, and quality of life. There is no consensus cut-off for the total sum fatigue score, yet, based on the Colombian normative data [44], we observed the mean values for each studied sub-group were all above 40, therefore we chose a clinical threshold of 40 for the total sum score.

### Machine Learning Analyses

Our machine learning analyses can be decomposed into three main steps: (1) the pre-training of the speech encoder model (See Figure 1b.) (2) the finetuning of machine learning models for each mental health aspect considered in this study (See Figure 1c.) and (3) extensive evaluations of the clinical threshold detection, selective detection, fairness assessments and severity estimations for each clinical scale (See Figure 1d. and Figure 2.).

**Figure 2.**
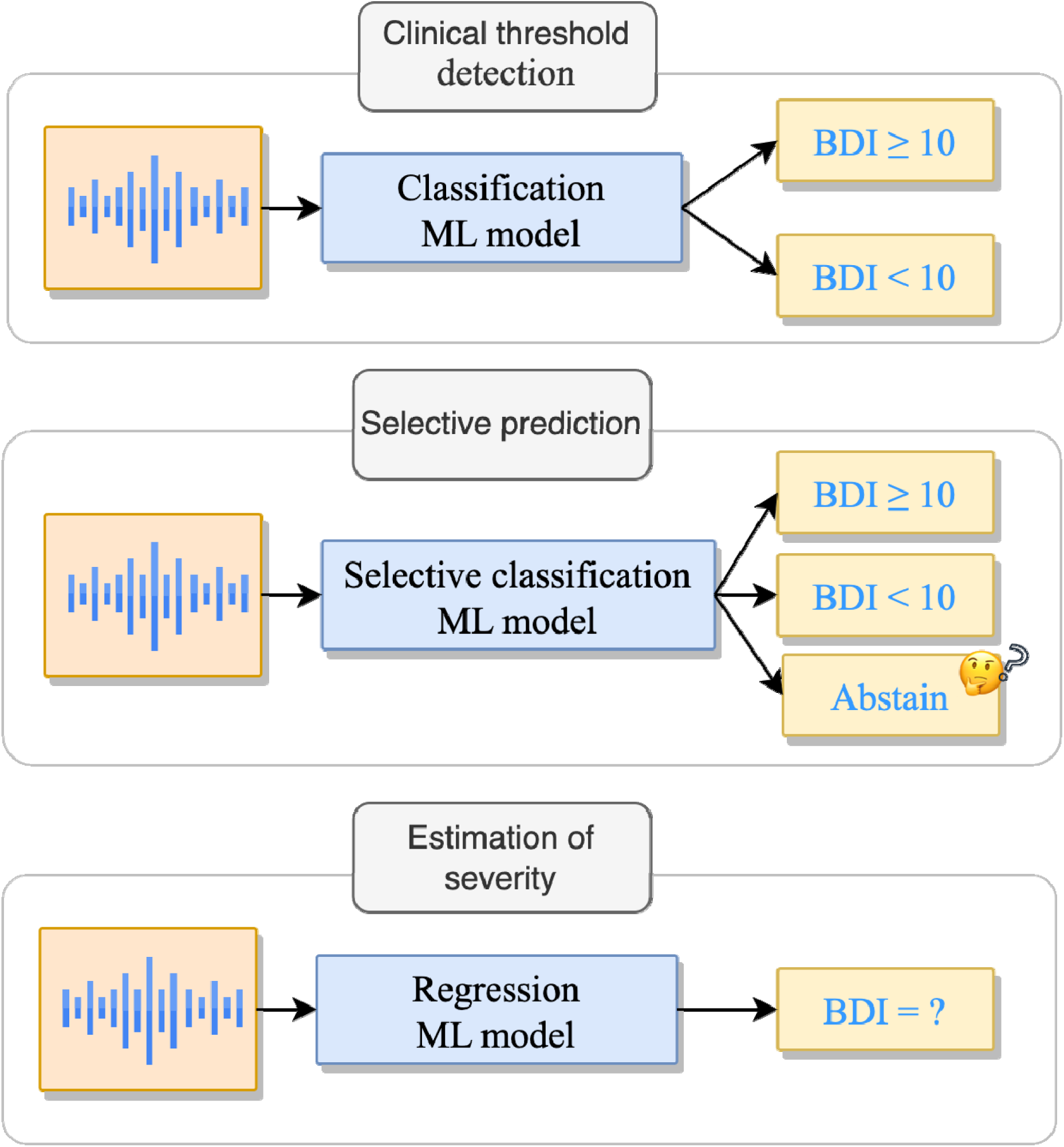
Schematic representation of the clinical tasks that are being assessed in this study. Each task is different in terms of the set and types of outputs. We illustrated the different tasks with the BDI clinical scale and its given threshold of 10.

#### Model pretraining and tuning

Audio intensity is normalized per sample, and we compared the three main approaches to obtain representations for large scale speech models: (1) Speaker recognition ia ThinResNet model with 34 layers, which takes as input speech samples encoded as 40-Mel Spectrograms with a hop length of 10ms and a hamming window. the same architecture ResNet as in [45], (2) we also considered a transformer [46] architecture adapted for speech, Hubert[47], trained in a self-supervision fashion, i.e. to predict masked neighbor embeddings; finally (3) we evaluated a transformer architecture Whisper pretained to tackle automatic speech recognition.

Speaker recognition as a pretraining task has proven great results in mental health and neurology [12, 48]. The ThinResNetis pre-trained on the VoxCeleb2 dataset [49], which is publicly available and contains over 1 million utterances from 6,112 speakers, from 145 nationalities. The VoxCeleb2 dataset consists of almost only continuous speech. The pre-training learning forces the model to organize speech in terms of speaker characteristics, as we illustrated above in the right panel of Figure 1b. We used an additive-margin softmax loss for this speaker identification task [50].

We also compared these models to a self-supervised model, HubertXL which exhibit great generalization for paralinguistic tasks such as emotion recognition[51] .

Hubert was trained on 960h of Librispeech [52]. Whisper is a recent robust Automatic Speech Recognition system based on a tranformer architecture trained on 680k hours of transcribed speech. Whisper training data is much bigger and more diverse, with noisy labels than other speech models. We considered three version of the model small, medium, and large (v3).

In this work, we did not fine-tune any of the speech encoder models on the Callyope-GP dataset, which we represented in Figure 1c. by a frozen speech encoder model.

For each speaker , we obtained a vector representation, a speech vector embedding denoted . We extracted segments of 20 seconds with 10 seconds overlap, and we compared different pooling of predictions, with mean and max pooling. Besides, as our data is imbalanced, we also compared classic sampling of examples to train models with undersampling of majority class, oversampling of minority classes. We used the default values of undersampling and oversampling: We found no differences with and without voice activity detection, so we used windowing for simplicity. Extraction was performed using Python (version 3.9) and the following packages were used to extract the acoustic features: pytorch 2.0.1, imbalanced learn, torchaudio 2.0.2, and voxceleb_trainer project [53].

For the finetuning for each task and each clinical score, we compared different machine learning algorithms on the validation set. For each task, once a model was selected, we retrained this final model on the concatenation of the training and validation sets and test on the held-out test, to avoid any inflated results. We used the scikit-learn implementation of each algorithm, splitting and evaluation [54].

For the speech collected in the Callyope-GP dataset, we applied the frozen speech encoder to each speech turn and propagated mental health assessment labels at the speech turn level, to train and compare the final machine learning model. At inference, for final evaluation, we pool predictions at the speaker level, after we obtained varying speech turns from a specific speaker.

We illustrated in Figure 2 each clinical endpoint, translated as a machine learning task based on the aforementioned procedure.

#### Clinical threshold detection (Classification)

We first compared the predictive power of the speech encoder to discriminate between individuals who are below or above the threshold for each clinical scale.

The distributions of positive and negative labels vary across clinical dimensions, and to take into account imbalance we reported the performances of the macro F1 scores, along the Area under the Curve (AUC) of the Receiver Operating Characteristic on the test set,.

We compared linear-based models (for classification Logistic Regression for classification with L2 regularization, Elasticnet linear model for regression), Tree-based models (Random forests with 100 estimators) and Gradient-boosting algorithms (Histogram-Based Gradient Boosting). Even though the pre-training phase captured information about the participants’ mental health, it is important to build a final model for each mental health dimension to be more specific and more sensitive. In ML terms, the mental health characteristics in speech are not necessarily linearly separable in the last vector space of the speech encoder.

#### Estimation of severity through predictions of individual scores (Regression)

The conventional approach in mental health assessment through speech analysis typically focuses on the group’s statistical analyses, or binary classifications of categorical outcomes, primarily discerning the presence or absence of a specific dimension. Yet, the risks of depression, anxiety, fatigue and insomnia exist along a spectrum of severity levels that exert varying degrees of influence on an individual’s well-being.

We go beyond traditional prediction and categorizations by integrating the estimation of severity through predictions of individual scores, employing regression machine learning models. This offers a more nuanced and comprehensive understanding of mental health dynamics, allowing for a more refined and personalized assessment. We evaluated our estimation of the severity of each total score with the Mean Absolute Error (MAE) between actual and predicted scores, and Pearson correlations (Table 4). MAE score directly measures how close the predicted scores are to the actual scores without considering the direction of error. We also reported the Pearson correlation and the P value between the actual test set and the predicted values.

### Fairness assessments: Quality-of-services for sex, age, and education level demographics

Machine learning systems can behave unfairly for different reasons and in multiple ways[28]. In medicine, the use of machine learning and predictive models should be carefully evaluated, especially for potential quality-of-service harms, i.e. it can occur when a system is not as performant for one specific group of people as it is for another group. We extensively studied the quality-of-service of harms for each clinical scale for the final predicted model, for each dimension: sex, age, and education level. We reported the Disparity Ratio (DR) based on clinical threshold detection F1 scores [57], to consider both false positives and false negatives and to be more stringent than equality of opportunity. The DR is computed as the fraction of the minimum F1 score across sub-groups divided by the average F1 score on the full test set: 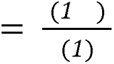 We did the same for pearson correlation for regression.

The higher the DR, the better it is as it means that the model performs equally well across groups, with the perfect DR being 1, i.e. each group has the same level of performance. We used the fairlearn toolkit to perform our fairness evaluations [58].

#### Selective clinical threshold detection (Selective prediction)

Machine learning approaches have made great strides in several domains, yet applications to high-stakes settings remain challenging. In our case, in mental health assessments, communicating appropriately the uncertainty associated with the system predictions is critical [59]. Yet, the communication of probabilities to human users is hard [60], and a pragmatic approach is to determine if an AI system is more likely to make erroneous predictions and defer these cases to clinicians. This setting can be cast as a selective prediction task for an AI system, the capability of a system to refrain from making a prediction when too uncertain (i.e., models saying “I don’t know”) [61, 62]. In this work, we followed the method from [63], we used the maximum output probabilities of the ML classification system as a way to measure uncertainty.

Based on a moving threshold we can obtain a specific ML system to choose to abstain when its output probabilities are too low. This specific ML system is evaluated based on the predictions it chooses to make only, thus there is a specific coverage and a specific accuracy, or risk. There is a natural tradeoff between the coverage of the ML system and its accuracy. Therefore, the way to evaluate a selective prediction task is the area under the curve for the risk-coverage curve.

## Results

### Data overview and demographics of participants

A total of 1150 participants were eligible and agreed to participate in our study. 865 completed the study, giving a recruitment yield of 75.2%. There was an equal split between Android (55%) and IOS (45%) devices used by participants.

The reasons for which participants were not included in this analysis were the following: one missing speech task, missing demographic information or one missing answer in the self-report questionnaires.

For our analyses, 605 participants were in the training set, 129 in the validation set, and 131 in the test set, and these groups did not differ in terms of demographics and mental health evaluations (cf Table 1). This yields a dataset sufficient (N>500) to evaluate error bars and predictive algorithms to avoid over-optimistic results [64]. Among the 865 participants, 275 (31.8%) were above the BDI screening threshold, and 146 (16.9%) were above the PHQ9 threshold, 133 (15.3%) above the GAD7 threshold, 371 (42.5%) above the AIS threshold, 489 (56.5%) above the MFI general fatigue threshold, 325 (37.5%) above the MFI physical fatigue threshold, 283 (32.7%) above the MFI reduced activity threshold, 379 (43.8%) above the MFI mental fatigue threshold, 209 (24.1%) above the MFI reduced motivation threshold, and 557 (64.4%) above the threshold of the total score of the MFI. We reported co-occurrences of people at risk for each dimension in the Multimedia Appendix 1.

**Table 1.**
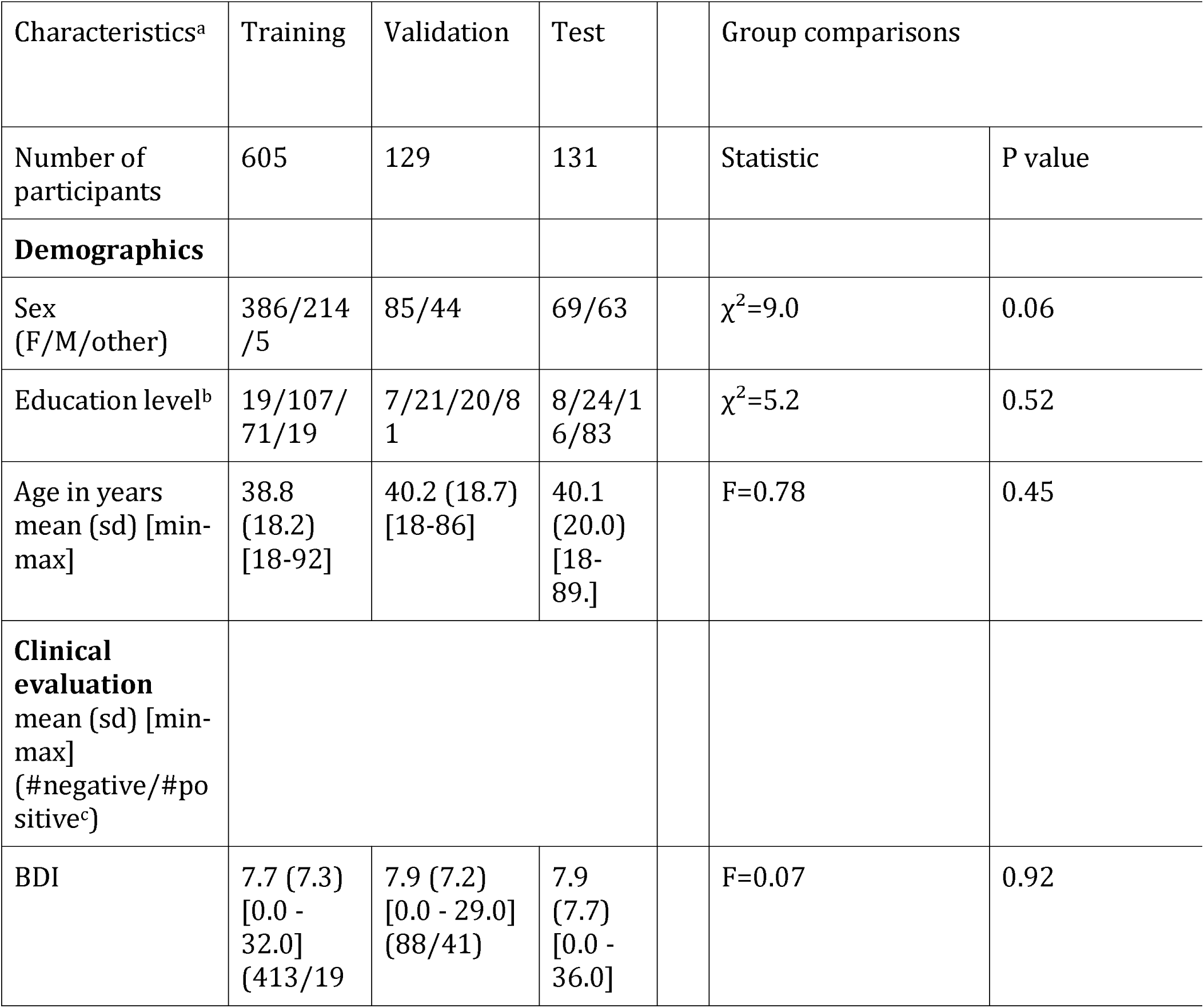
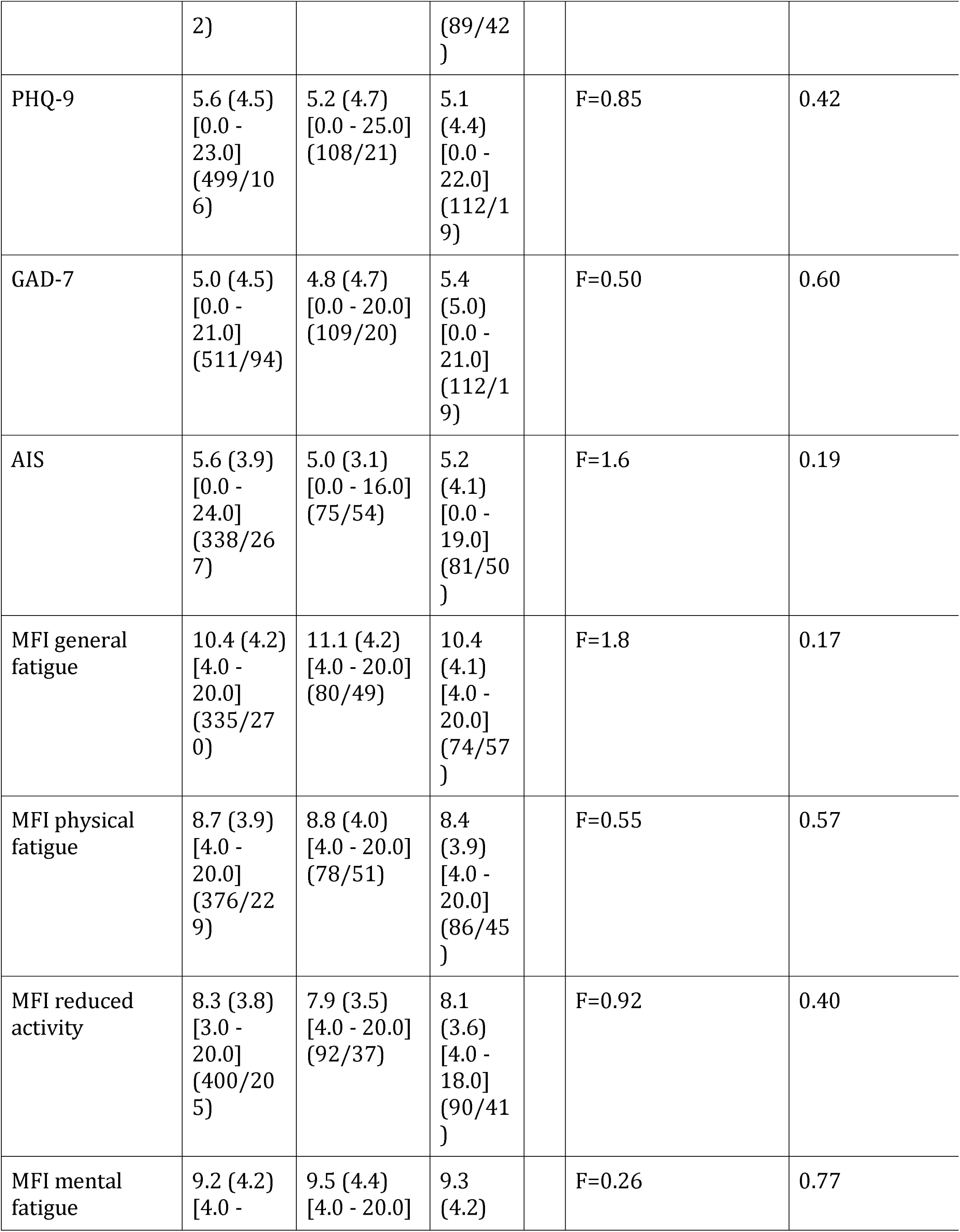
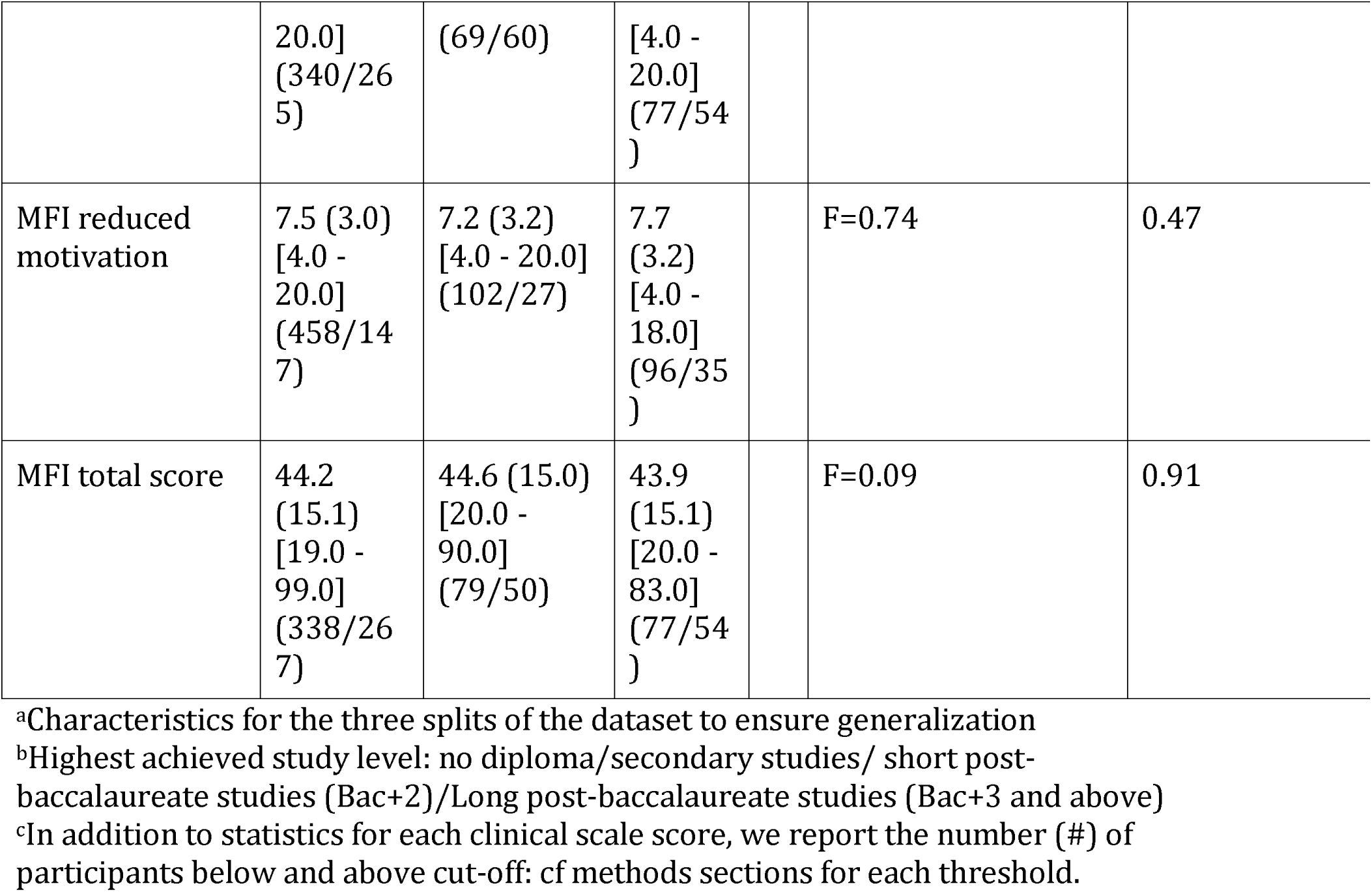
Demographic characteristics of the training, validation and test groups^a-e^. Categorical variables are compared with Pearson’s χ² test and one-way ANOVA for continuous variables. PHQ-9: Patient Health Questionnaire, BDI: Beck Depression Inventory, GAD-7: General Anxiety Disorder 7-item scale, AIS: Athens Insomnia Scale, MFI: Multidimensional Fatigue Inventory

### Machine Learning Analyses

We compared models and pipelines with the results on the validation set (See Table 5 in Appendix). Overall, all the Whisper models all outperformed other approaches, (WhisperM being the best with mean F1 score=0.56), while the speaker model performed the worst, even though still performing well. We found out that the performance of the pooling with the maximum prediction always outperformed the mean pooling, and undersampling and oversampling was helping. We found out that the linear-based models was outperforming random forest and gradient boosting on the validation set. Thus, we retrained linear algorithms, with max pooling and the WhisperM frozen speech encoder on the combination of the training and validation sets and reported the final results on the held-out test set in Figure 3, Figure 4, Table 2, Table 3 and Table 4 and Appendix Figure 2.

**Figure 3.**
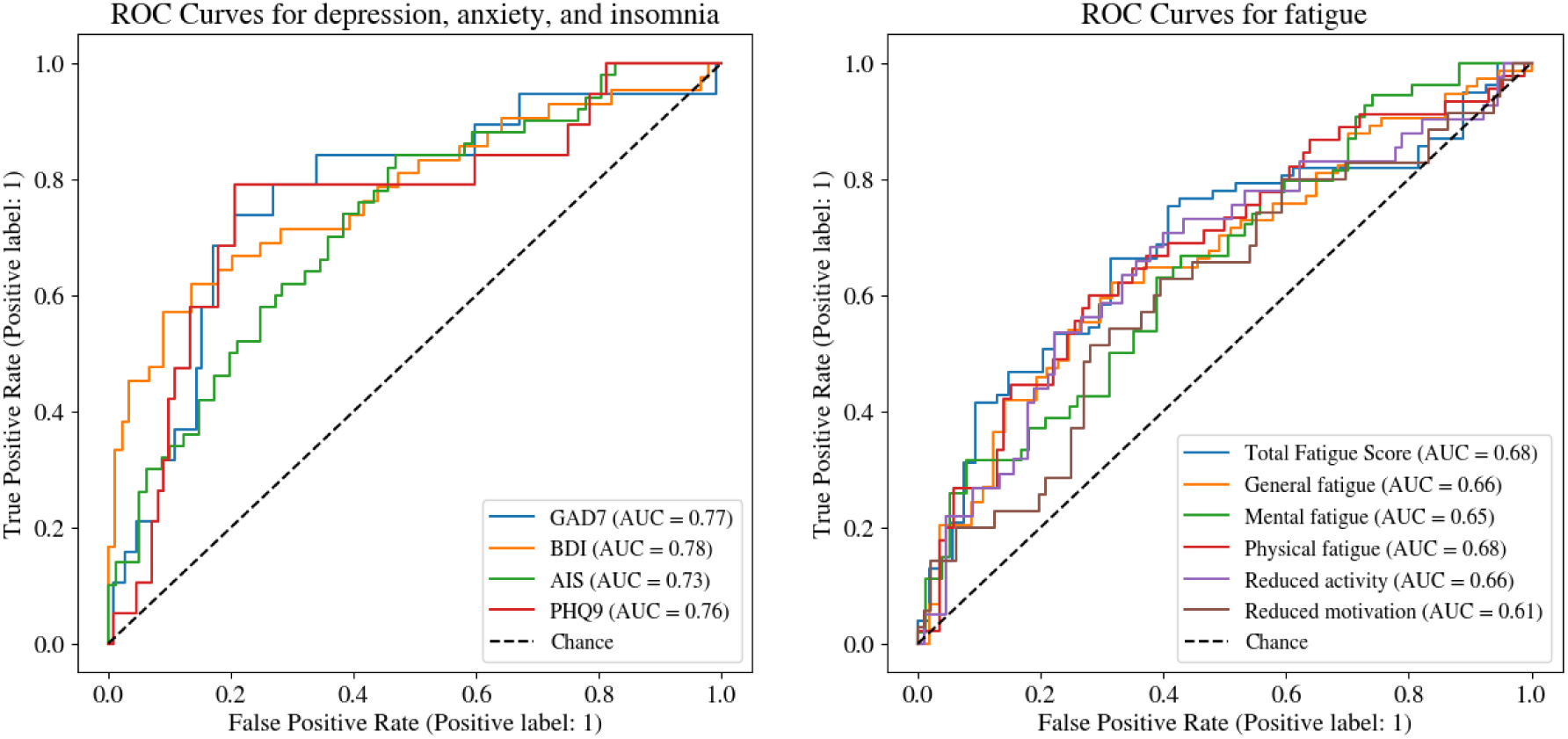
ROC curves for the clinical threshold detection task on the held-out test set. (Left panel) ROC curves to detect clinically relevant thresholds for depression (PHQ-9, BDI), anxiety (GAD-7), and insomnia (AIS). (Right panel): ROC curves to detect clinically relevant thresholds for fatigue components (MFI), general fatigue, physical fatigue, reduced activity, mental fatigue, reduced motivation and total fatigue. PHQ-9: Patient Health Questionnaire, BDI: Beck Depression Inventory, GAD-7: General Anxiety Disorder 7-item scale, AIS: Athens Insomnia Scale, MFI: Multidimensional Fatigue Inventory.

**Figure 4.**
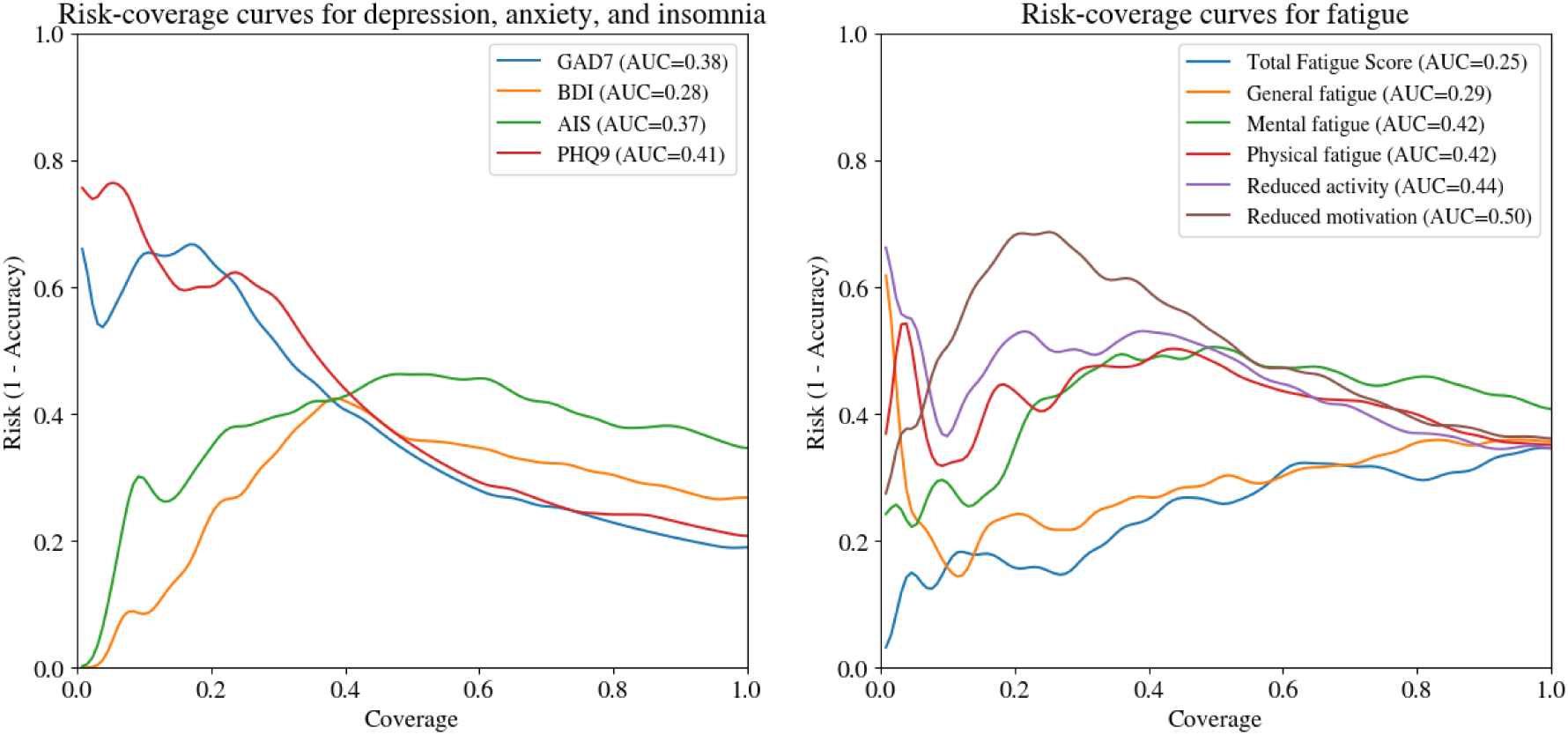
Risk-Coverage curves on the held-out test set for the selective clinical threshold detection task on the held-out test set illustrate the models’ selective screening ability, i.e., risk detection capabilities with the ability to abstain when too uncertain. Curves are smoothed for clarity with a gaussian blur, but not used to compute AUC. A lower AUC is better, and 0 is the perfect score. (Left panel) Risk-Coverage curves selectively detect clinically relevant thresholds for depression (PHQ-9, BDI), anxiety (GAD-7), and insomnia (AIS). (Right panel): Risk-Coverage curves to detect clinically relevant thresholds for fatigue components (MFI), general fatigue, physical fatigue, reduced activity, mental fatigue, reduced motivation and total fatigue. PHQ-9: Patient Health Questionnaire, BDI: Beck Depression Inventory, GAD-7: General Anxiety Disorder 7-item scale, AIS: Athens Insomnia Scale, MFI: Multidimensional Fatigue Inventory.

**Table 2.**
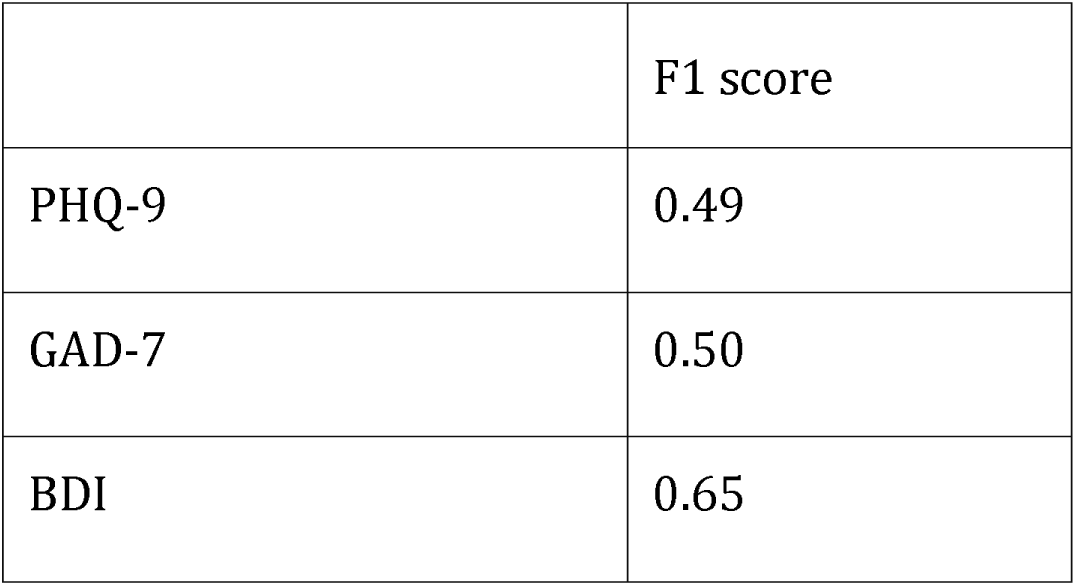
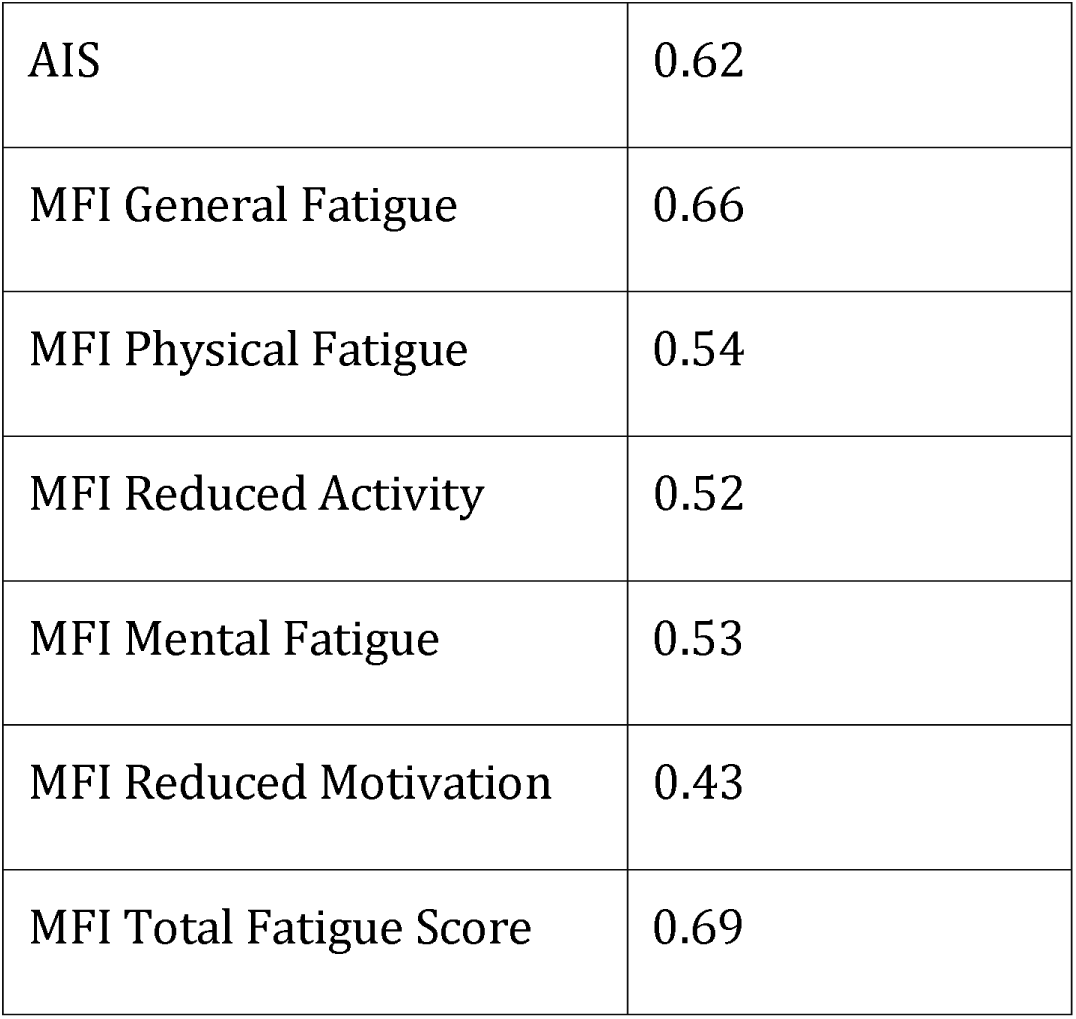
Estimation for clinical threshold detection severity results on the held-out test set for the different considered dimensions of mental health (Classification). Higher F1 is better, and 1 is perfect. PHQ-9: Patient Health Questionnaire, BDI: Beck Depression Inventory, GAD-7: General Anxiety Disorder 7-item scale, AIS: Athens Insomnia Scale, MFI: Multidimensional Fatigue Inventory.

**Table 3.**
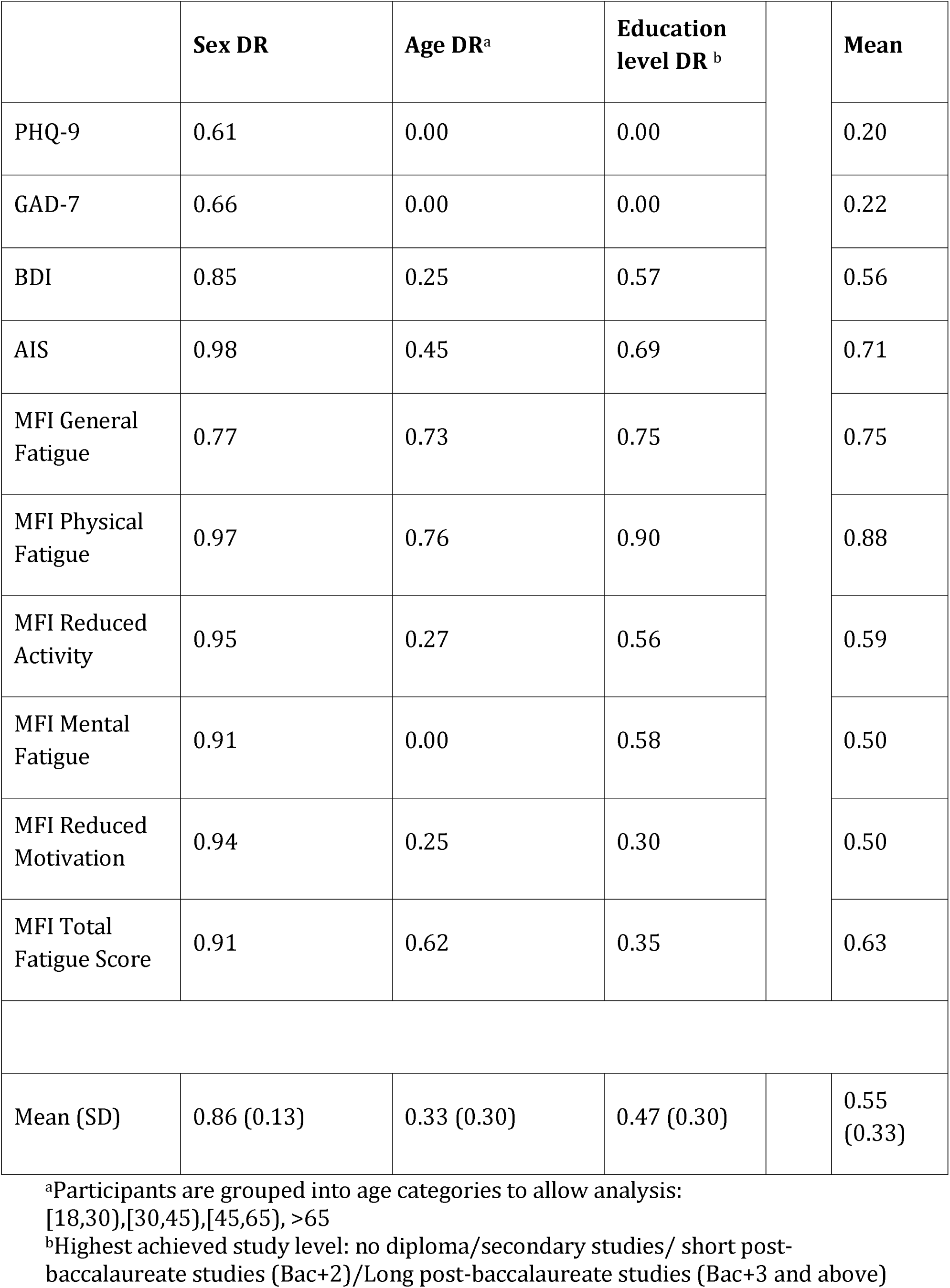
Disparity ratios (DRs) based on the F1 scores for sex, age and education levels to assess fairness, on the held-out test set, for the clinical threshold detection (Classification) for the different considered dimensions of mental health. Higher DR is better, and 1 is perfect DR. Mean value per score is reported. Mean and Standard Deviation are reported per sensitive dimension. PHQ-9: Patient Health Questionnaire, BDI: Beck Depression Inventory, GAD-7: General Anxiety Disorder 7-item scale, AIS: Athens Insomnia Scale, MFI: Multidimensional Fatigue Inventory.

**Table 4.** Estimation of severity results on the held-out test set for the different considered dimensions of mental health (Regression). We reported Mean Absolute Errors (MAE) and Pearson correlations between actual and predicted values. Lower MAE is better, and 0 is perfect. A higher Pearson correlation is better, +1 is perfect. All correlations were significant P <1×10^-14^. PHQ-9: Patient Health Questionnaire, BDI: Beck Depression Inventory, GAD-7: General Anxiety Disorder 7-item scale, AIS: Athens Insomnia Scale, MFI: Multidimensional Fatigue Inventory.

|  | **Pearson $r$** | **MAE** |
| --- | --- | --- |
| PHQ-9 | 0.47 | 3.1 |
| GAD-7 | 0.48 | 3.2 |
| BDI | 0.49 | 4.9 |
| AIS | 0.43 | 2.9 |
| MFI General Fatigue | 0.38 | 3.3 |
| MFI Physical Fatigue | 0.32 | 3.0 |
| MFI Reduced Activity | 0.31 | 2.9 |
| MFI Mental Fatigue | 0.34 | 3.1 |
| MFI Reduced Motivation | 0.32 | 2.5 |
| MFI Total Fatigue Score | 0.44 | 11.3 |

**Table 5.** Results on the validation set for the clinical threshold detection task on the validation set for the F1 score and AUC score, averaged across clinical scales. We displayed the top 3 approach for each model. We underlined the best approach per frozen speech model, and bolded the best overall.

| Frozen speech model | Pooling | Sampling | Classifier | F1 score |
| --- | --- | --- | --- | --- |
| Random classifier (Dummy) | n/a | n/a | n/a | 0.15 |
| Speaker Recognition ResNetSE34 | max | undersample | Logistic Regression | 0.518 |
|  | max | undersample | Random Forest | 0.514 |
|  | max | oversample | Logistic Regression | 0.510 |
| Self-supervised HubertXL | max | oversample | Logistic Regression | 0.532 |
|  | max | undersample | Logistic Regression | 0.528 |
|  | max | undersample | Gradient boosting | 0.522 |
| WhisperS | max | oversample | Logistic Regression | 0.535 |
|  | max | undersample | Logistic Regression | 0.522 |
|  | max | None | Logistic Regression | 0.513 |
| WhisperM | max | oversample | Logistic Regression | 0.560 |
|  | max | oversample | Gradient boosting | 0.547 |
|  | max | undersample | Logistic Regression | 0.542 |
| WhisperL | max | oversample | Logistic Regression | 0.548 |
|  | max | undersample | Logistic Regression | 0.545 |
|  | max | undersample | Gradient boosting | 0.527 |

#### Clinical threshold detection (Classification)

The clinical threshold detection performed well based on the speech data and our developed system (Figure 3 and Table 2). All systems outperformed the chance levels ). Based on the AUC the classification results were the highest for the BDI score (AUC=0.78, F1=0.65) and based on the F1, it was the Total MFI score(AUC=0.68, F1=0.69) and the lowest for both metrics the MFI reduced motivation (AUC=0.61, F1=0.43).

### Fairness assessments: Quality-of-services for sex, age, and education level for classification

We computed the sensitive attributes DRs (sex, age, and education level) and assessed the differences in the quality of the c made by the speech-based system (Table 3) for classification. Overall, the classfication speech-based system had a better quality-of-service for sex (mean 0.86, SD 0.13), and the worst was for age (mean 0.33, SD 0.30). We also identified that the detection of PHQ9-7 had the worst quality-of-service disparity (Mean of DRs = 0.20, and the best quality-of-service was obtained with the score AIS (Mean of DRs = 0.71), the MFI General Fatigue (Mean of DRs = 0.75), and the MFI Physical Fatigue (Mean of DRs = 0.88). We also observed that only the MFI General Fatigue and MFI Physical Fatigue obtained a good performance for age (MFI General Fatigue Age DR=0.73; MFI Physical Fatigue Age DR=0.76), and, except for PHQ-9 and GAD-7, all DRs were satisfactory or high for sex.

#### Selective clinical threshold detection (Selective prediction)

We also evaluated the capabilities of speech-based models to selectively predict the different clinical thresholds. We observed great performances for BDI, AIS and MFI General Fatigue (Figure 4). The model that selectively predicts the risk of depression based on the BDI score achieved the best result (BDI risk-coverage AUC=0.28). The other scores could not achieve such feats of important coverage with no risk.

#### Estimation of severity through predictions of individual scores (Regression)

We reported the regression results for the estimation of severity in Table 3. Speech-based models obtained significant results for all clinical variables, based on the evaluation with the Pearson correlations (all P <1×10^-14^). The strongest correlations between the prediction and the actual scores on the held-out test were found for BDI score (r=0.49), GAD7(r=0.48) and the PHQ-9 (r=0.47). The lowest correlation was found for the MFI Reduced Activity (r=0.31).

Speech-based models also obtained great results in terms of absolute errors. We observed less than 3 points of MAE for AIS and reduced activity. All other scores were predicted on average with less than 5 points, except for the MFI Total Fatigue Score, since its range is [13–88].

## Discussions

We aimed to explore the full capabilities and limitations of using speech data extracted from 865 participants in the general population to predict the presence or absence of different mental health self-reported symptoms: depression, anxiety, insomnia and the different dimensions of fatigue.

We built a fully automated speech-based machine learning system that takes as input the audio waveform collected from one simple speech task performed on our smartphone application. The models were trained and calibrated on training and validation sets of participants, and we demonstrated the system’s generalization on the held-out test set of 131 participants.

The results indicated that ML-based systems using speech only as input could identify participants above clinical thresholds for depression, insomnia, total fatigue components, but to a lesser extent anxiety and fatigue sub components. All classification results were above chance levels for each clinical threshold.

This result was confirmed with an extensive fairness analysis of quality-of-service for age, sex and education levels. Depression, insomnia and different dimensions of fatigue clinical threshold detection results were particularly consistent for sex, slightly less for age and to a lesser extent for education level. Anxiety risk identification fell behind in accuracy overall and was also unequal per group. The extension of our clinical threshold detection system to be able to abstain, with selective prediction, was conclusive, even for anxiety. Risk-coverage areas under the curves remained low, for insomnia, total fatigue, and depression detection through BDI. Finally, we showed that speech-based models could also predict the severity, with prediction of exact scores, moving beyond binary interpretations of score thresholds. All correlations between the predicted scores and the actual scores given by participants were significant, exhibiting strengths ranging from 0.31 to 0.49.

Our study builds upon existing mental health research on speech analysis and extends the insights for deployment into clinical practice. For risk and anxiety depression in the general population, we found similar strong performances such as [26, 27, 65, 66]. Our recruitment and involvement of participants was in person. For medium-sized datasets with face-to-face recruitment (below 1000 participants, such as ours), it can be observed that crowd-sourced recruitment of participants via online platforms needs more training data to yield the same level of model accuracy [65, 67]. This was also observed in a large study, with over 6000 participants, with online data recruitment for risk detection in the general population in American English [66]. This discrepancy could be explained by the fact that data quality is variable on web crowdsourcing platforming, especially for participants’ psychiatric evaluations [68], and also voice recording through laptops.

Our study revealed discrepancies in both clinical threshold detection and estimation of severity through self-reported depression scores between BDI and PHQ-9. This underlines the inherent limitations of score conversion and the crucial role of individual-level assessment in capturing the nuanced and different expressions of depression [35]. This reinforces the necessity of developing assessment tools and interpreting results with meticulous attention to individual variability, particularly by scrutinizing model performance at the individual level, mirroring real-world clinical scenarios.

Our study uniquely addresses the co-occurrence of perceived fatigue and reported insomnia, both prevalent mental health concerns, which could be detected simultaneously through speech analysis. While prior research, like the work by [20], has explored how sleep deprivation impacts specific vocal features like prosody and voice quality, no previous study has delved into the combined influence of fatigue and insomnia on speech. Addressing this gap is crucial because these conditions often co-occur and significantly impact the symptom trajectory and potential development of other mental health issues [69]. The prevention of recurrent sleep problems can prevent other mental health troubles or relapses. The observed co-occurrence of symptoms, particularly insomnia with other clinical dimensions, highlights the interconnected nature of these symptoms and syndromes.

Finally, , to the best of our knowledge, this is the first study to assess the fairness and selective prediction capabilities in speech-based mental health assessments. This is of prime importance since speech signals can be heavily influenced by a multitude of factors such as age, sex, weight, and height [70]. Age factor was the least preserved in our grouped performances, this can be attributed to change in voice changes due to hormones [71], and normal aging affects the different parts of the vocal production system: larynx, respiratory system, resonators, saliva system and the individual’s global emotional status [72]. In addition to lower performances compared to other mental health dimensions, the anxiety risk detection performance collapsed for certain groups of demographics. This could be explained by the heterogeneity and low positive examples in our Callyope-GP dataset. Even though there are limits concerning some groups of individuals, selective classification offers an option to potentially remediate these variable quality-of-services, ensuring a deployment in clinical settings and still bringing overall clinical utility.

### Limitations

While valuable, this study has some limitations. The French monolingual, medium-sized dataset (300<N<1000) needs more diverse data to achieve better generalizability, and the non-longitudinal design misses insights on symptom evolution. Besides, the use of self-assessment scales introduces a potential bias because they rely heavily on the insight of participants. A limitation of our study is the use of a fixed train-dev-test split. This split approach, while convenient for comparing results across different tasks, can introduce bias and limit the generalizability of our findings. Future research with larger samples, longitudinal designs, and the inclusion of pathological data is crucial for exploiting the full potential of voice biomarkers in mental health. Studying various and diverse speech tasks and prompts also holds the potential for multiple benefits related to user adherence. Indeed, tailored tasks can address specific mental health needs, cater to individual preferences and boost engagement. Varying prompts can reduce user fatigue and sustain interest, leading to more consistent system use and richer data collection.

## Conclusions

In conclusion, this study demonstrates the potential of speech-based systems for detecting and predicting various mental health symptoms in the general population. While challenges remain regarding real-world applications and ensuring fairness across the population demographics, our findings pave the way for further development and responsible integration of such tools into clinical settings, advancing personalized mental health assessment and intervention. In future work, we will extend this study by including longitudinal data, adding more diverse linguistic and geographic data, and including more severely affected patients who are already followed by mental health practitioners. We will also look into fairness and uncertainty mitigation methods to improve the performance of our systems.

## Data Availability

The participants of this study did not give written consent for their data to be shared publicly, so due to the sensitive nature of the research supporting data is not available.

## Acknowledgements

The authors are thankful to all the participants who volunteered for this research study. Without their active involvement, the present study would not have been possible. We also would like to thank each of the speech pathology interns who helped with the subject recruitment and made sure that the protocol was completed successfully.

## Conflicts of Interest

RR, XN, AL, MdG, MD, and AB are shareholders of Callyope and VO was a former employee of Callyope.

## Abbreviations

JMIR: Journal of Medical Internet Research
RCT: randomized controlled trial
ML: Machine Learning
PHQ-9: Patient Health Questionnaire
BDI: Beck Depression Inventory
GAD-7: General Anxiety Disorder 7-item scale
AIS: Athens Insomnia Scale
MFI: Multidimensional Fatigue Inventory.
DR: Disparity Ratio to measure fair quality-of-service
n.s.: Non significant
MAE: Mean Absolute Error
PR: Precision-Recall curve
ROC: Receiver Operating Characteristic curve
AUC: Area Under the Curve

## Multimedia Appendix 1

**Figure Appendix 1.**
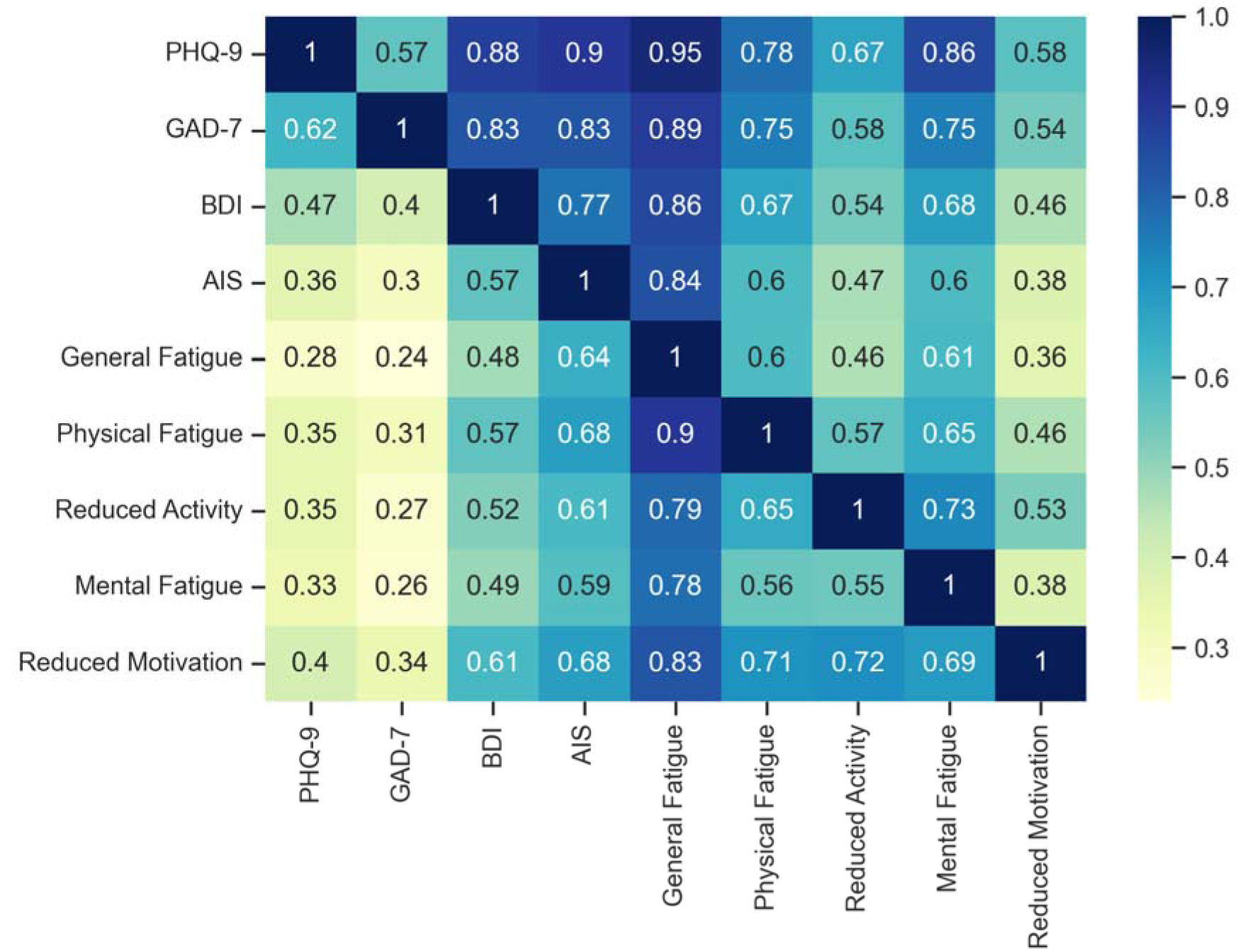
Co-occurrences percentage of people at risk for each dimension, normalization is done per row: first row can be read as 57% of individuals who are at risk for depression based on the PHQ-9 are at risk of GAD-7.

**Figure Appendix 2.** ROC curves for the clinical threshold detection task on the held-out test set. (Left panel) PR curves to detect clinically relevant thresholds for depression (PHQ-9, BDI), anxiety (GAD-7), and insomnia (AIS). (Right panel): ROC curves to detect clinically relevant thresholds for fatigue components (MFI), general fatigue, physical fatigue, reduced activity, mental fatigue, reduced motivation and total fatigue. PHQ-9: Patient Health Questionnaire, BDI: Beck Depression Inventory, GAD-7: General Anxiety Disorder 7-item scale, AIS: Athens Insomnia Scale, MFI: Multidimensional Fatigue Inventory.

## Notes

### Competing Interest Statement

RR, XN, AL, MdG, MD, and AB are shareholders of Callyope and VO was an employee of Callyope.

### Funding Statement

This study was funded by Callyope.

### Author Declarations

After fully understanding the explanation of the study, all participants signed a consent form to participate in the study, in line with the Declaration of Helsinki, current Good Clinical Practice guidelines, and local laws and regulations. All procedures were approved by the French National Institutional Review Board, (identifier 23.00748.OOO2L7#I for the Committee for the Protection of Persons).

### Summary of Updates

Many updates due to code base checks, modelling changes, results changed and review updates taken into account.

